# Compliant citizens, defiant rebels or neither? Exploring changing COVID-19 vaccine attitudes and decisions in Bradford, UK: Findings from a follow-up qualitative study

**DOI:** 10.1101/2022.06.24.22276852

**Authors:** Bridget Lockyer, Rachael H Moss, Charlotte Endacott, Shahid Islam, Laura Sheard, the Bradford Institute for Health Research Covid-19 Scientific Advisory Group

## Abstract

**Background:** COVID-19 vaccines have been the central pillar of the public health response to the pandemic, intended to enable us to ‘live with Covid’. It is important to understand COVID-19 vaccines attitudes and decisions in order to maximise uptake through an empathetic lens.

**Objective:** To explore the factors that influenced people’s COVID-19 vaccines decisions and how attitudes towards the vaccines had changed in an eventful year.

**Design and participants:** This is a follow up study that took place in Bradford, UK one year after the original study, between October 2021 and January 2022. In-depth phone interviews were conducted with 12 (of the 20 originally interviewed) people from different ethnic groups and areas of Bradford. Reflexive thematic analysis was conducted.

**Results:** 11 of the 12 participants interviewed had received both doses of the COVID-19 vaccine and most intended to have a booster dose. Participants described a variety of reasons why they had decided to have the vaccines, including: feeling at increased risk at work; protecting family and others in their communities, unrestricted travel and being influenced by the vaccine decisions of family, friends and colleagues. All participants discussed ongoing interaction with COVID-19 misinformation and for some this meant they were uneasy about their decision to have the vaccine. They described feeling overloaded by and disengaged from COVID-19 information, which they often found contradictory and some felt mistrustful of the UK government’s motives and decisions during the pandemic.

**Conclusions:** The majority of participants had managed to navigate an overwhelming amount of circulating COVID-19 misinformation and chosen to have two or more COVID-19 vaccines, even if they had been previously said they were unsure. However, these decisions were complicated, and demonstrate the continuum of vaccine hesitancy and acceptance. This follow up study underlines that vaccine attitudes are changeable and contextual.

**Patient or Public Contribution:** The original study was developed through a rapid community and stakeholder engagement process in 2020. Discussion with the Bradford Council Public Health team and the public through the Bradford COVID-19 Community Insights Group was undertaken in 2021 to identify important priorities for this follow up study.

## Introduction

Worldwide, the COVID-19 pandemic has brought a multitude of immediate challenges, including severe illness and deaths, periods of social restriction and isolation, national lockdowns, travel restrictions and extreme economic disruption. However, since the identification of several successful COVID-19 vaccines at the end of 2020, countries across the globe have attempted mass vaccination programmes in order to bring the COVID-19 pandemic ‘under control’.^1^

In the UK, and specifically in England, the Government has considered vaccination to be the central pillar of the public health response to the pandemic, intended to enable us to ‘live with Covid’.^2^ Since the Pfizer/BioNTech vaccine was approved for use in the UK on 2nd December 2020, a large-scale COVID-19 vaccination programme was introduced and rolled out at a rapid pace. By 17th June 2021, all adults over the age of 18 had been given the opportunity to receive their first (of two) COVID-19 vaccine(s). Several months later, a booster (third dose) was introduced, and all adults in England were given the opportunity to book an appointment to receive a COVID-19 booster vaccine by the end of 2021. By the 12^th^ June 2022, 90.9% of over 12s in the UK had received their first dose, 86.3% had received their second and 71.9% had received their booster or third dose^3^ (See Figure 1 for key COVID-19 milestones in England). Studies focused on COVID-19 vaccine acceptance found that people were motivated by wanting to protect themselves and others, by trust in science and the evidence of vaccines’ effectiveness, by trust in their GP and by wanting to get back to normal life after periods of social restrictions. ^4-6^

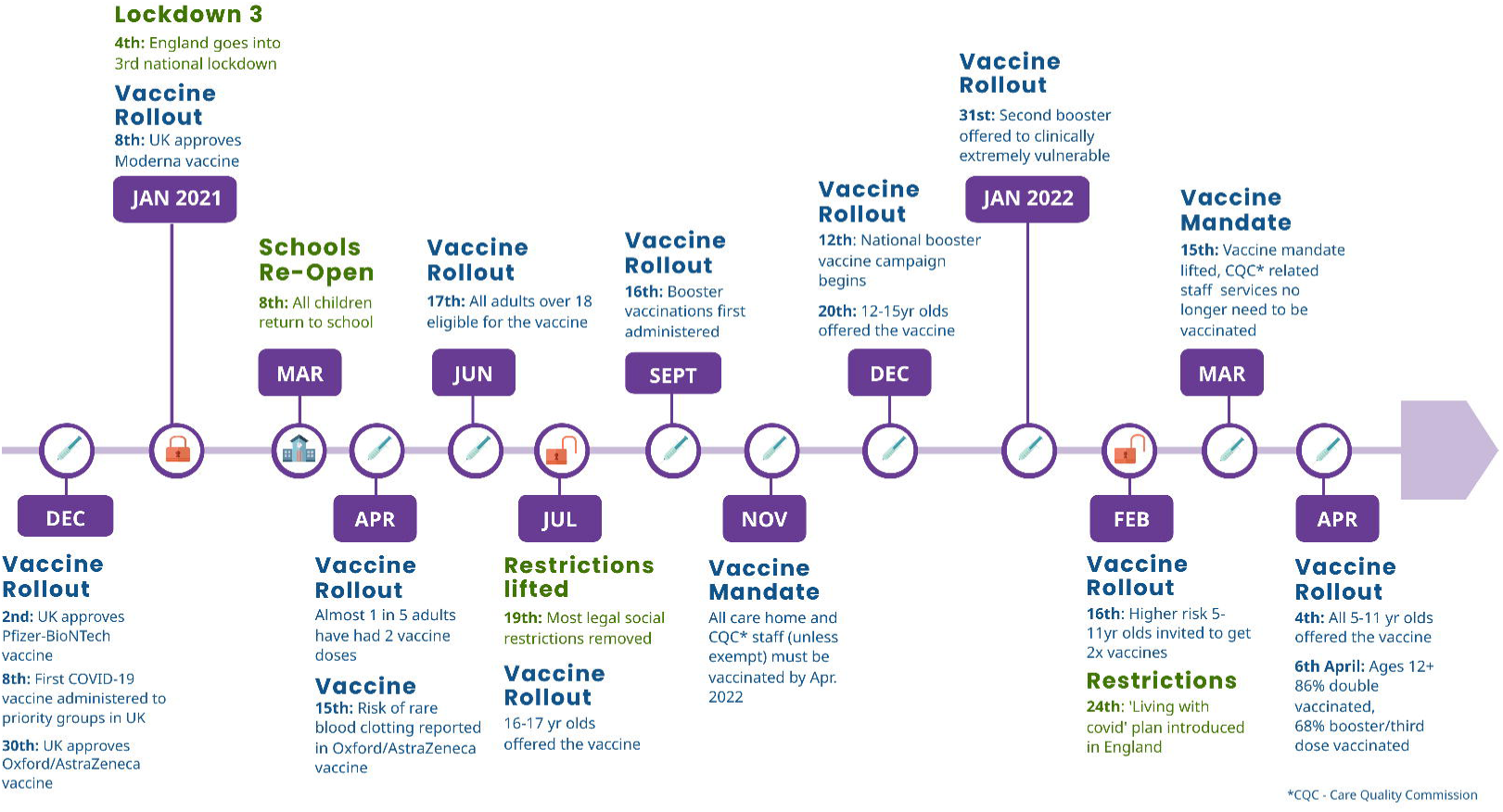

Before the introduction of COVID-19 vaccines, there were concerns about vaccine hesitancy and unequal uptake, due to historical patterns of low vaccine uptake.^7^ In the UK, there were early indications that some population groups were more hesitant than others. Higher vaccine hesitancy was associated with women, people from younger age groups, lower education levels and being from certain minority ethnic groups.^8-10^ This hesitancy appears to have been somewhat borne out in the uptake figures. Vaccine uptake was found to be lowest in some of the communities for whom COVID-19 has the biggest risk, including the Pakistani, Black Caribbean, Black African and Bangladeshi communities, undocumented migrants, and studies have found a strong negative association between socioeconomic deprivation and the rate of declining COVID-19 vaccinations.^11-15^ Conversely, higher vaccine acceptance was associated with being White British, older and more educated. ^14^ Since the COVID-19 vaccines were first rolled out there have been serious concerns about widening health inequalities as a result of uneven uptake and there is emerging evidence to suggest that is the case.^16, 17^

Much of the recent public discourse around COVID-19 vaccines has been divisive, and has often strayed into racist and classist territory to explain patterns of uptake amongst different population groups.^18, 19^ Vaccine hesitancy, which refers to a delay in acceptance or refusal of vaccination despite its availability^20^, is in itself a contested term and considered by some to place blame on certain population groups or individuals, when wider structural forces are at play.^21-22^ These include access to and relationship with health services, long standing mistrust in institutions, poor or inappropriate methods of health communication and socio-economic factors such as being unable to travel to vaccination centres or get time off work to attend a vaccination appointment. However, it is our understanding that using the term ‘vaccine hesitancy’ can and should take these factors into account. In 2014, the SAGE Working Group on Vaccine Hesitancy developed the confidence, complacency, convenience model of vaccine hesitancy, the 3C model.^20^ This model highlights: 1) confidence and trust barriers, 2) complacency and perception of risk barriers and 3) convenience, structural and socio-economic barriers. Conversely, greater vaccine acceptance is associated with greater trust in health authorities,^23,24^ greater availability of accurate and accessible information for those considering vaccines^25,26^ 26 and confidence in vaccine effectiveness and length of disease protection.^27^

Where we think the term ‘vaccine hesitancy’ can be useful is that it is not binary, people exist on a hesitancy and acceptance spectrum or continuum.^20^ Whilst the spectrum ranges from full acceptance to total refusal, there are a lot of people in the middle and their beliefs, situations and decisions can alter and shift, moving them up and or down the spectrum.

Our previous study explored the impact of the COVID-19 ‘infodemic’ and ‘misinfodemic’ ^28,29^ through interviews with 20 citizens in Bradford, UK. We found the deluge of conflicting, alarming and often inaccurate health information intensified feelings of confusion, distress and mistrust, leading to greater uncertainty about whether to have the vaccine. This study was conducted in Autumn 2020 before any COVID-19 vaccines were approved for use, so questions about vaccination intention were hypothetical. Subsequent studies have enabled us to further understand that exposure to misinformation, particularly online, increases vaccine hesitancy.^31,32^ However, this process did not take place in a vacuum and existing levels of trust in governments, media, science and the health service were found to be an important influence, and lower levels of trust were found in ethnic minority groups due to long-standing institutional racism.^33-36, 14^ Attempts to counter misinformation have been mixed; providing clear communications on the risks and benefits of the vaccine alone was found not to be sufficient in increasing vaccine uptake nor was correcting.^37,38^ Many studies have recommended local approaches, offering accessible information for different groups in the community and leveraging trusted in-group messengers.^13,29,39^ On a structural level, increasing trust in health organisations, science and information via reputable sources is key but remains challenging.^29,40^

This study follows up our 2020 study, returning to conduct interviews with the same participants. ^30^ Unlike other recent studies, which have largely and understandably focused on the reasons why some people choose to get vaccinated and why some people choose not to, we explore vaccine motivations, hesitancy and acceptance on a spectrum. As we had interviewed the participants before, we had a good understanding of their COVID-19 beliefs, experiences and vaccine intentions, and therefore could consider if, how and why they have changed. This also allows us to start to explore the implications of sustained exposure to COVID-19 information and misinformation on both those that have chosen to have the vaccine and those who have not.

## Methods

### Study design and PPI

This follow up descriptive, inductive qualitative study was completed as part of a larger mixed-method, longitudinal research study to provide actionable intelligence to local decision makers, developed in response to community and stakeholder consultation processes described in our previous article. ^30^ Our earlier findings were shared and discussed with Bradford public health teams and with the public through a COVID-19 Community Insights Group. This group was established in March 2021, and meets every 6 weeks to engage with residents from the district to understand how they and their communities are coping during the COVID-19 pandemic. The group consists of 14 (well-connected and diverse) community members. During the development of a follow up study, we went back to both the public health team and the community groups to ask them what their priorities were at that time and the topics they thought we should include on the interview schedule so these could be incorporated. We used in-depth interviews to explore the same individuals’ health experiences and beliefs during COVID-19, one year on. University ethical approval for the follow up study was secured in October 2021.

### Study setting

Our follow up study was conducted in Bradford, a city in the North of England. Bradford and its surrounding district is the fifth largest metropolitan district in England and is an area of high deprivation and ethnic diversity, with established Pakistani, Bangladeshi, Eastern European communities. Since March 2020, Bradford has experienced a relatively high number of COVID-19 cases, and stricter lockdown measures from July 2020 which remained in place until the introduction of the tier system in October 2020.^30^ This pattern has continued throughout subsequent infection waves. High rates of COVID-19 in areas like Bradford are likely to be due to greater deprivation, high population density and a higher than average number of multi-generational households.^30^

### Sampling and data collection

We returned to the same participants from our previous study, contacting them via email and phone. We attempted to contact all 20 previous participants, two declined to be interviewed on a first approach, four agreed to be interviewed but were repeatedly unavailable (which we took to be a subtle decline) and we were unable to reach two.

Fieldwork took place between November 2021 and February 2022. Six women and six men participated; their ages ranged from 25 to 86 years old, but two thirds were aged between 35 and 54. In terms of ethnicity, participants identified as Asian or Asian British (Pakistani, Indian and Bangladeshi) (n=5), White British (n=4) and White Other (Eastern European) (n=3) (see Table 1 for participant demographics). The participants lived in five different Bradford postcodes, representing some degree of variation in geography and deprivation status. Many of the participants were in paid or volunteer community roles; other jobs included teacher, supermarket worker and childminder.

**Table 1:**
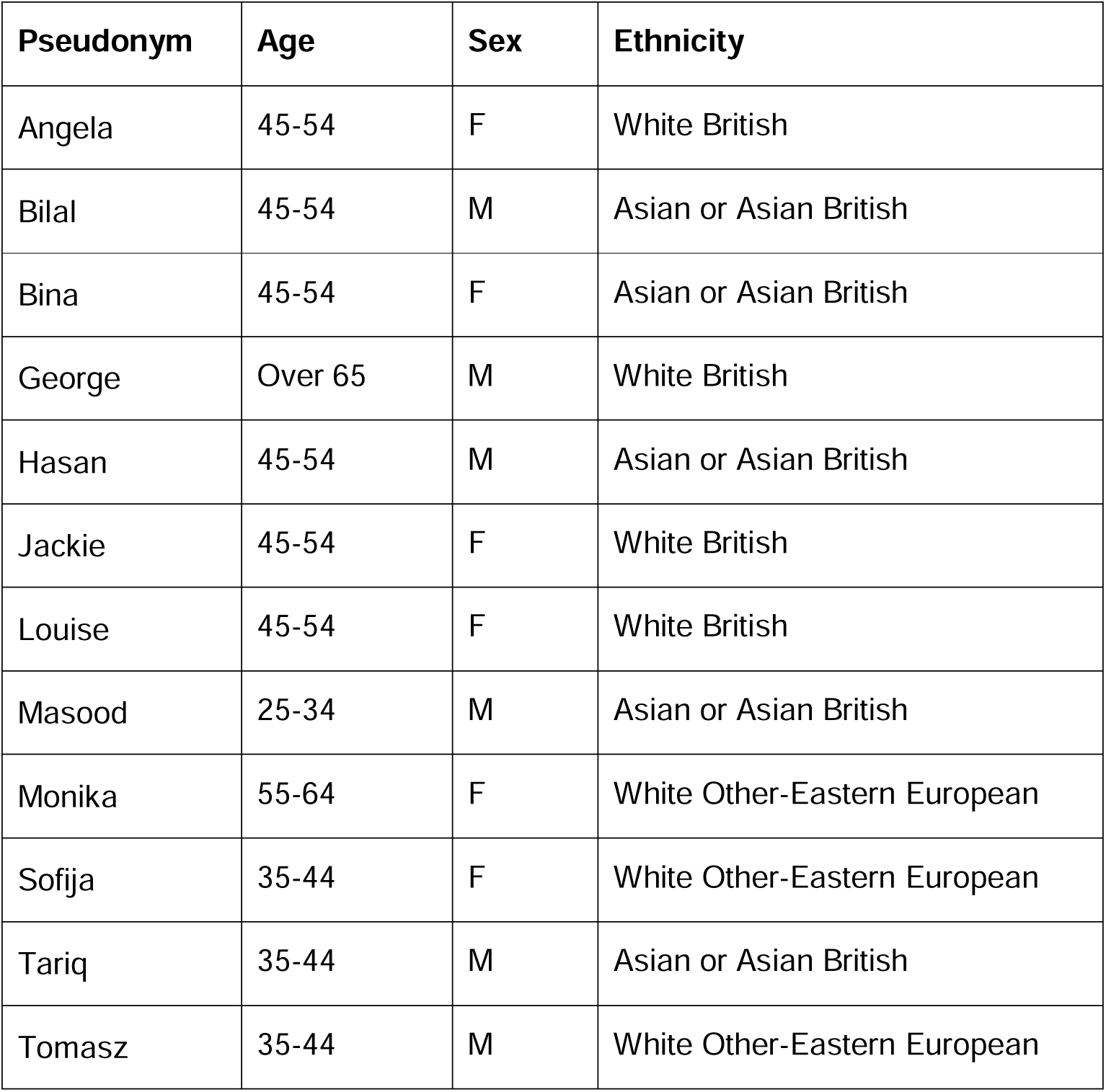
Participant Demographics.

All interviews were conducted in English over the phone by the first author. The interviews ranged in duration from 25 to 120 minutes, with the average length being around 50 minutes. All participants gave written, informed consent through one of the following methods: (a) emailing a completed consent form or, (b) emailing/texting stating that they had read the information sheet and consent form and fully consented to taking part in the study. In addition, all participants confirmed consent verbally at the start of each interview. All interviews were digitally recorded and transcribed by a professional transcriber with identifying information removed and participants’ names pseudonymised.

### Interview questioning

Headline topic guide questioning was adapted from the previous guide, updated to take into account the current COVID-19 vaccine programme. For example, in this guide we included questions about participants’ intentions to have the COVID-19 booster vaccine. Questions were added through consultation with local public health teams and the community group described above. The format of the topic guide and interview questioning was flexible to allow participants to voice what they considered important.

### Analysis

We undertook the analysis using the principles of reflexive thematic analysis.^41^ All transcripts and interview field notes were coded independently by the first, second and third authors. We held two analysis sessions to identify commonalities and differences in the interview narratives and worked towards ordering the data into loose themes. These themes were then refined by the first three authors with example quotes. The first author subsequently analysed all interviews and conducted further interpretive work to write up the findings, sense checking with the other authors as necessary. The analysis was conducted manually without the use of a software package. The analysis was wholly inductive, and, as such, we did not structure it on any existing theoretical frameworks and it was not based on the themes developed in our previous study.

## Findings

Out of the 12 people interviewed for this follow-up study, 11 had had at least two COVID-19 vaccines, and most were intending to get their booster. In the original study, 4 people in this follow up group (Jackie, Louise, Tariq and Sofija) had been unsure and hesitant about whether they would have the vaccine when it became available to them. The findings are presented in two sections. The first explores reasons why the participants and their friends and family had the vaccine, which included travel, protecting family and friends, influence of others and personal experience of COVID-19. The second section ‘consequences of the (mis) infodemic’ builds on our previous work which contended that misinformation had stoked confusion, mistrust and distress during the pandemic, increasing participants’ hesitancy about a COVID-19 vaccine. In this follow up study, we found that although most participants had chosen to be vaccinated, continued interactions with misinformation and feeling overwhelmed by conflicting information about COVID-19 generally, had led to feelings of uneasiness about the vaccines’ effects, overload and disengagement with health information and a sustained sense of mistrust in government.

### Why did people get vaccinated?

Participants described various reasons why they and their friends and family had chosen to have COVID-19 vaccine(s). People were largely concerned about their health and the health of those they were closest to. Participants who had jobs which put them and their family at greater risk of virus transmission, such as Masood who worked as security guard, Louise who worked in a supermarket and Angela who worked as childminder, said that this prompted them to get a COVID-19 vaccine as soon as it became available to them:

> *‘I work like face-to-face like with the customers so I can understand because my family is at home, so I just…straight away I get it [the vaccine]’ (Masood)*.
>
> *‘I mean I did it for my son basically, you know, who’s been quite poorly as a baby with asthma and I didn’t want, I work, you know, I won’t say I work on the frontline but I am a key worker and I, I work on a checkout in a supermarket so I could potentially be bringing Covid home from my workplace without knowing about it’ (Louise)*.
>
> *My daughter (aged 17) didn’t want it. She weren’t keen on it. I don’t know, she didn’t give an excuse but I didn’t give her a choice. I told her she had to because we work with children and children are the carriers of a lot of germs and that’ (Angela)*.

For some, this sense of protecting others extended to their wider community and society:

> *‘I did it for the greater good, I went and had mine done’ (Jackie)*.

A factor which appeared to influence participants’ vaccine decisions was that they now had had more personal experience of COVID-19. In the original study conducted in Autumn 2020 none of the participants had knowingly had COVID-19, this time four had been ill with COVID-19 and the majority had a close family member or friend who had had the virus. Angela discussed her husband’s experience of the virus, which was particularly worrying as he had existing respiratory issues:

> *‘Me husband did, he got really poorly. Well, he nearly died. He couldn’t breathe but he has a breathing machine so if I sent him into hospital they’d only put him on that so I got him on his breathing machine’ (Angela)*.

Sofija discussed her step-son who also had existing health conditions and was ill for months:

> *‘[He was] very bad, and very difficult to fight with this, and I think he’s still got like a loss of taste and smell, so it took a long time, he recovered’ (Sofija)*.

Another of the main motivations for having COVID-19 vaccines appeared to be practicality, as the vaccines enabled individuals to travel to different countries more freely:

> *‘I had a battle with my own children. We had to talk it through. They were talking nonsense but they are so fickle as young people as soon as they said “you can’t go on holiday until you’ve had your vaccines”, they all had their vaccines done’ (Bina)*.
>
> *‘One friend who was sceptical about it, after talking to us has now had the vaccine. But I think many people in the Polish community are taking it now because they want to travel’ (Tomasz)*.

Some people described having the vaccine as common sense and ‘an easy decision’ (Monika). A few participants suggested that their previous life experience and having had many vaccines in the past made them less concerned about having a COVID-19 vaccine. This included Robert who had been in the armed forces and as such, was required to have a lot of vaccinations as part of his work:

> *‘No, as I say I’ve had so many vaccines and injections and inoculations over the years from everything from cholera, yellow fever, you name it, I’ve had it’ (Robert)*.

Bina, although younger than Robert, described coming from an age group which valued vaccines, and how this caused her to be more ready, informed and willing to get a COVID-19 vaccine than subsequent generations:

> *‘I am a believer, I’m a generation of vaccines, I was vaccinated from an early age…*.*So I believe in vaccine, I understand the purpose of vaccines…’ (Bina)*.

Interestingly, despite their very willing acceptance of the COVID-19 vaccine, Robert and Bina had declined their annual influenza vaccine invitations, believing that it was not necessary for them because they were healthy. Whilst they discussed why they had the COVID-19 through the lens of social responsibility, they appeared to view the flu vaccine as a more personal health choice.

Some participants did suggest that they were initially worried about the immediate side effects and perceived long term health effects of the vaccine, but felt reassured when they saw and heard about people they knew having it:

> *‘*…*when you see more people getting the vaccine then it kind of like gives you a bit more faith…it’s okay to get it done’ (Louise)*.

As seen in some of the responses above, the influence and active encouragement of friends, family and peers to get the vaccine was evident. Sofija described how her husband, who was initially worried about the vaccines’ side effects, came to the decision to have it four months after it became available to him:

> *‘[Seeing] work colleagues they have been, they’re all vaccinated and so I think this pushed him as well to see that they’re fine, yeah’ (Sofija)*.

She also described appealing to his emotions, and asking him what would happen to their children if they were both ill or died from COVID-19. At the time of interview, she said her husband was trying to persuade his brother to get the vaccine.

## Consequences of the (mis)Infodemic

### Uneasiness about vaccine safety

Despite their decision to have the vaccine, participants still conveyed some uneasiness about the long-term impacts of COVID-19 vaccines:

> *‘I don’t know what it’ll do to my body in the future, I don’t know if there’ll be any […]I’ve no idea what could go wrong. I weighed it up and it might, it might not…and no, I just needed to get it done, needed to do my bit’ (Jackie)*.

Jackie’s account suggested she had struggled to weigh up what was best for her and what was best for society. She indicated that she thought that her long term health had been put at risk by having these vaccines. Similarly, Louise said that although she had been persuaded to have the COVID-19 vaccines by seeing her friends who were nurses have it, she still harboured thoughts that the vaccine she received could have unknown long-term health consequences:

> *‘So, you know, and I think when you see your nurse friends and family, you know, having the vaccine it gives you a bit more confidence that, you know, if, well if they’re going to get wiped out I’ll go with them [laughs]. You know? I mean if, if we’ve all had this vaccine and we’re going to die in the next couple of years we’ll all go together, won’t we?’ (Louise)*.

The possibility that the COVID-19 vaccines could have lasting health implications appeared to be rooted in misinformation that the participants had encountered. All participants showed an awareness of misinformation about COVID-19 vaccines within the interviews. As in the previous study, most were keen to make clear that they did not believe any ‘conspiracy’ stories about the vaccines, yet common tropes about the vaccines’ safety, such as them changing your DNA or causing infertility, were part of their narratives:

> *‘They did say that it changed DNA, one of them, didn’t they and stuff?’ (Angela)*.
>
> *‘I’ve read there is a special code and the code will change your DNA and you will die. And younger people wouldn’t be able to have children’ (Sofija)*.
>
> *‘My daughter said she didn’t want to take a vaccine because what if they affected the time when she wanted to have children…When the story came out that the Pfizer vaccine was affecting the foetal part of the body when they’re pregnant and so on she was then reading up on how people had been affected’ (Bina)*.

### Overload and disengagement

Whilst the spread of misinformation may have begun online and been facilitated by social media, it was evident that it was being discussed amongst friends and families and had influenced people’s beliefs and decisions. Tomasz discussed why a friend of his had refused to have the vaccine:

> *‘I think it’s because he’s got you know, other friends around yeah, and maybe there’s a lot more people who are sceptical about it in his circles’ (Tomasz)*.

Tariq, the only participant not yet vaccinated, had been heavily influenced by his friends:

> *‘Some friends, that I’ve made over last couple of years they’ve got quite close to me and they’re like “oh you’d better not take it, you’d better not take it, you don’t know what’s in it…you’re not well already and who’s going to look after you if something happens to you?’ (Tariq)*.

Tariq was saturated in COVID-19 misinformation. His friends had invited him to join a very active WhatsApp group which was inundating him with negative videos and pictures about the safety and inefficacy of the vaccine. He discussed some of this content:

> *‘Like some people have said “oh this person’s had the jab and then they’ve got, they’ve been paralysed or something’s happened to them or they’ve got ringing in their ears, you know, they’ve had a heart attack or they’ve had to go and have more surgery or kidney’s failed or they’ve had a blood clot”, stuff like that. So I don’t know if it’s true. Did the British Airways pilots die of blood clots or was that just a hoax?’ (Tariq)*.

Other stories he discussed were the building of a camp in the UK to imprison people who refused to get the vaccine and the local hospital hiring actors to make it look full of COVID-19 patients. He was obviously conflicted by the information he saw and was not sure what to believe:

> *‘I’m trying to stay away from it now because it’s just like, it’s proper confusing and I’m trying my best to stay away from it but it’s quite hard because I’ve not left the group and I’m thinking “shall I leave the group? Shall I leave the group or not?” and I’ve not done and then I’m still listening to what’s coming through and I’m like thinking “I should have left the group, I should have left the group”. But then you’re thinking like “hang on, are they making sense?”‘ (Tariq)*.

Tariq’s sense of confusion and feeling of being overwhelmed was very apparent. Although he was the only one who had not yet had the vaccine, other participants described being overwhelmed and confused about COVID-19 information and misinformation, and indicated these feelings had led them to take less notice of the news:

> *‘I turn off the TV when I see people arguing about vaccinations’ (Jackie). ‘I get fed up of hearing it, I turn the telly off’ (Angela)*.

Making sense of the deluge of information about COVID-19 since March 2020 appeared to have had an emotional and mental toll, which led to people purposefully choosing to disengage:

> *‘I’ve avoided the news and social media, I made a decision to because of my health anxiety’ (Monika)*.
>
> *‘I’m just keeping abreast of information but not letting it take over my life like it did at the beginning…’ (Bina)*.
>
> *‘I feel better without this information…because I’m very emotional’ (Sofija)*.

The distress Sofija experienced from seeing both information and misinformation during the early part of the pandemic was still clearly influencing her ability to engage:

> *‘They scared me so much that yeah [laughs], that you know, that yeah, depression and, yeah, it was horrible. I don’t want to be in that state anymore, that’s why I’m like, I don’t want to hear anything because it affected me so much’ (Sofija)*.

There was some evidence that this disengagement had led to people being less informed about what the current official guidance on vaccines was. When Tomasz was asked about whether he was planning to get his booster jab he did not appear to have considered it much, although he was soon to be eligible and it was being encouraged amongst his age group:

> *‘If we need to take a booster vaccine then I’ll take it. And the virus has, you know, mutated and stuff like that, so I think it’s worth, you know, taking but I will read into that. Actually, that one I might check-out you know, how it works and*…*’ (Tomasz)*.

The information people appeared to take most account of was local information, provided by the council and local health centres:

> *‘So I signed up to Bradford Council email, so I get emails, updates quite regularly from Bradford Council and they’re quite good…I think that it’s been really good. I work with a community centre and they get information obviously from the Council and they’ve found the information really, really good’ (Bilal)*.

### Mistrust

One of the reasons why people described feeling overwhelmed and confused was because they were not sure who to trust. Official health information felt contradictory, and at the time of the interviews, there had been several reports about those in the Government not abiding by lockdown rules. Sofija said she had found information from the Government:

> *‘…very mixed. The politicians not following their own messages and you know, it’s not like trustworthy’ (Sofija)*.

There was a lot of scepticism about the Government’s response to the pandemic, with people thinking that they have been scaremongering and lying about the numbers of deaths related to COVID-19:

> *‘I’ve seen them on telly and you don’t know if they’re telling truth or making it worse. Saying thousands and thousands have died and it’s going here and going there. No, you don’t know if they’re just doing it to scaremonger the whole of the world’ (Angela)*.

Tomasz felt the Government had allowed private companies to ‘monetise the pandemic’ and said a lot of his friends felt more strongly than he did:

> *‘I think lots of my friends don’t trust yeah, the Government yeah. And then people are generally fed-up with all the information. I see lots of comment about oh stop scaremongering blah, blah blah, something like that, you know. So, people I think are fed-up, they don’t want to hear about it’ (Tomasz)*.

For Hasan, despite believing in COVID-19, laughing at ‘conspiracy theories’ and having had two vaccines and booster when offered as it is better to be ‘safe than sorry’, he was incredibly cynical about the Government’s response and felt they had exaggerated COVID-19’s impact for political and financial gain:

> *‘I’m probably the wrong person but for me Covid has just been an excuse and a reason for government in order to do certain things and be able to do things, so it’s been a God send and that’s all it is at the end of the day’ (Hasan)*.

Ultimately, mistrust had not prevented our participants from getting the vaccine but it was easy to understand from their responses the ways in which it was contributing to hesitancy.

11 of the 12 participants in this follow up had chosen to have at least two COVID-19 vaccines and most had or were planning to get their boosters. However, their decisions and beliefs were complex and there was still uneasiness, disengagement and mistrust. Some of those within the 2020 study that had been most hesitant (such as Jackie and Louise) remained so, even though they had opted to have two vaccines. Sofija was relatively hesitant in 2020, anxious about the vaccine’s safety, but was now much more accepting, even encouraging family and friends to take it. Participants like Angela and Hasan who said they were going to have the vaccine in 2020 and did, still suggested that they believed some wider conspiracy theories about COVID-19. Bina and Robert were very positive about the vaccine in 2020 and 2021/22 but this acceptance did not lead to an overarching acceptance of all vaccines, as they were firm that they did not want a seasonal flu vaccine. Tariq, our one participant still unvaccinated, appeared to be even more conflicted and overwhelmed than he had been in 2020.

## Discussion

This follow up study aimed to understand people’s COVID-19 vaccine decisions and explore their health beliefs and experiences around COVID-19 and COVID-19 vaccines. We found that most participants had chosen to have two or more COVID-19 vaccines. Their reasons and the reasons of those close to them included: protecting their health and the health of loved ones; travelling without restrictions; closer experience of COVID-19 and the positive influence of others around them having the vaccine. These reasons corresponded to the findings of other studies which explored vaccine acceptance, although the influence of others was more pronounced and there was more ambivalence about the vaccines’ safety and effectiveness. ^4-6^

In our previous study we found that exposure to COVID-19 misinformation had led to confusion, distress and mistrust and was contributing to uncertainty in vaccine intentions. Our follow up study found that the majority of participants and those close to them had managed to overcome the tidal wave of misinformation and get the vaccine. However their accounts still suggested exposure to and engagement with misinformation, particularly around vaccine safety and long term effects, which was causing some to be uneasy about their decision. In addition, after over 18 months of interacting with distressing, confusing and often contradictory COVID-19 information and misinformation in news, social media and within their social circles, they were feeling overwhelmed. As a result, several participants described actively disengaging with anything related to COVID-19 in order to feel more in control and less anxious. Many were distrustful and suspicious about the Government’s response to the pandemic, some were even doubting what they had been told about the severity of the COVID-19 virus.

Our findings illustrate the continuum of vaccine hesitancy and acceptance, and recognises that those who have chosen to have the vaccine may still have doubts and concerns. This is important to understand because it means that we cannot take their current vaccine acceptance for granted, it is based on current social and political conditions and patterns which can change.^42^ This aligns with the study of expected vaccine uptake,^43^ which found that both conditional acceptance and rejection of COVID-19 vaccines were dynamic and volatile. Through following up with the same participants, we were able to see that people’s location along the hesitancy/acceptance spectrum is changeable. Sofija and her husband were good examples of that, moving from vaccine hesitancy and delay to encouraging others to get their vaccines. This is further evidence that people who appear “vaccine hesitant” can indeed be convinced.^43^

This method has also allowed us to understand the different and compounding reasons why people may move further along the spectrum towards acceptance despite reservations, including health concerns, peer/family encouragement and practicalities. Our study has also highlighted that people’s levels of vaccine acceptance can differ for different vaccines. For example, some participants were happy to have the COVID-19 vaccine and were not willing to have a seasonal flu vaccine. Being vaccinated for COVID-19 was deemed to be socially responsible, but being vaccinated for flu was a personal health choice, suggesting that people did not fully understand the social and human costs of a widespread flu outbreak. This reminds us that vaccine hesitancy and acceptance should be looked at in the historical, political and socio-cultural context in which vaccination occurs.^44^ We had less insight into the beliefs and motivations of those still refusing to be vaccinated as this two wave study depended on participants agreeing to be re-interviewed. Yet the internal struggles of Tariq and his willingness to discuss them demonstrated that even those who appear to be staunchly resistant could have the potential to move towards acceptance within a less confusing and distressing environment.

Often local and national media write-ups of studies which report and discuss COVID-19 vaccine uptake amongst different population groups (including our own work) have been met with social media comments which contain a ‘them and us’ narrative, sometimes with implicit and explicit racist and classist tropes. Lower than average vaccine uptake in Bradford has been blamed by online commentators on the Pakistani and Eastern European populations, the poor and the less educated.^45^ Public discourse on both ‘sides’ have called one another stupid and uneducated, for either their perceived compliance or defiance. ^45^ Yet our participants’ views and decisions were nuanced and complex, and largely understanding of others’ doubts and fears, because they shared some of them too. This study adds further weight to a need to move away from the binary of vaccine hesitancy and acceptance, not only because it can contribute to stigmatising certain demographic groups, but because it is incorrect and unhelpful.

## Implications for Policy and Practice

We found misinformation, whether it was regarded as misinformation or not, to still be present and sometimes influential in the experiences and narrative of our participants. Numerous studies have advocated for local, targeted, community-driven and accessible health information,^13,29,39^ and we think this study gives further weight to persisting with this, particularly because trust in national government was low. The quality of local government-public relationships is positively associated with pro-vaccine outcomes, including more frequent risk information seeking, pro-vaccine attitudes, and greater vaccination intention^46^ and our study found that participants’ most trusted health source was the local council. Although the misinformation machine is global, continuing to foster and develop strong and trusting relationships locally can help erode some of its impacts.

This study also found that encouragement and positive discussion about COVID-19 vaccines with family and friends was persuasive. Seeing vaccine acceptance normalised amongst friends, colleagues and acquaintances was also influential, as people trusted the decisions of those close to them. This further underlines the importance of health messaging that leverages personal relationships and positive emotions.^47^ Our findings were able to capture how overwhelmed people were by the COVID-19 (mis)infodemic, and the potential for disengagement with future COVID-19 booster(s) or other vaccine campaigns as a result. Again, clear and positive public health communication should be prioritised over messaging that is likely to engender further anxiety and distress.

### Limitations

As this was a follow up study our sample size was necessarily restrained. In addition, we found that some of those who said they were very unlikely to have a COVID-19 vaccine, were unwilling to talk to us this time or were unreachable. This is perhaps understandable given how politicised and fraught vaccine decisions have become. As a result, we have less insight into the uptake and beliefs of some of those who appeared most hesitant in 2020.

## Conclusions

The public discourse around COVID-19, and vaccines in particular, has often felt polarised. There has been an assumption that people have either been compliant citizens or defiant rebels, assumptions which often have classist and racist undertones. The findings of this study ask us to consider the feelings and behaviours of the vast majority of the population who are neither. By following up with the same group of people from a largely deprived and multi-ethnic city, we could appreciate, in context, how and why they made their decisions and more deeply explore the complex influences of family and peers, health (mis)information and (mis)trust in institutions. The majority of the participants had chosen to be vaccinated, but this was not without some uneasiness and their narratives still contained threads of misinformation and mistrust. As well as underlining the persistent effects of misinformation, this study re-emphasises vaccine hesitancy/acceptance as a continuum, rather than as a binary concept. In doing so, we hope to contribute to a greater and more empathetic understanding of what shapes the health beliefs and behaviours of all of us on the continuum.

## Data Availability

The data that supports the findings of this study are available on reasonable request from the corresponding author. The data are not publicly available due to privacy or ethical restrictions.

